# The Impact of 6-Month ART Dispensing (6MMD) on Retention in Malawi’s HIV Program: A Target Trial Emulation Study

**DOI:** 10.64898/2026.02.16.26346393

**Authors:** Khumbo Shumba, Idah Mokhele, Elizabeth Kachingwe, Lise Jamieson, Matthew P. Fox, Sydney Rosen, Timothy Tchereni, Stanley Ngoma, Sophie Pascoe, Amy N. Huber

## Abstract

**Background:** Six multi-month dispensing (6MMD) of antiretroviral therapy (ART) for HIV treatment clients has expanded rapidly in the past decade, but its effect on individual outcomes in routine (non-trial) care is still poorly documented and based on observational programmatic data. Malawi launched 6MMD in April 2019 and scaled-up implementation in 2020. We compared retention in care for clients who received 6MMD to those who did not using a target trial emulation (TTE) approach to minimize bias.

**Methods:** We used routine clinical data from Malawi’s Electronic Medical Record (EMR) system to identify ART clients eligible for 6MMD in 27 districts from 01/2020-12/2021. Eligible participants were non-pregnant adults (≥18 years), on ART for ≥6 months, clinically stable as evidenced by a dispensing duration of 3 months (3MMD), and with no prior 6MMD exposure. We created four six-month trials, defined eligibility at the start of each trial period, and classified participants as either receiving 6MMD or non-6MMD (dispensing duration of 1-3 months) within the six-month interval. Follow-up started at 6MMD enrollment for the 6MMD arm or the first visit in the trial enrollment period for the non-6MMD arm. Retention at 12 and 24 months was defined as having a clinic visit within 12-24 (trial 1-4) and 24-36 (trial 1-2) months from trial enrollment. Using an intention-to-treat approach, we estimated adjusted risk differences (aRD) with 95% confidence intervals (CI) using a Poisson regression model with an identity link function and robust standard errors adjusting for age, sex, duration on ART, facility type, regional location, WHO clinical stage at ART initiation. Pooled RDs were estimated by accounting for within-subject variation in a Poisson regression model using data from all trials.

**Results:** Of the 159,801 unique patients eligible for this study (65% female, median age 37 years), 74% (118,910) were ever enrolled in 6MMD. Retention rates at 12 months (trials 1-4) and 24 months (trials 1-2) were consistently higher in the 6MMD group than the non-6MMD group. The pooled risk for retention was 3% higher in the 6MMD vs non-6MMD groups (aRD 3.0%; 95% CI: 2.8%-3.3%) at 12 months and 2.0% higher (aRD: 2.0%; 95% CI: 1.7%-2.4%) at 24 months.

**Conclusions:** We observed slightly higher retention in care rates in Malawi at 12 and 24 months among patients on 6MMD compared to those receiving shorter medication dispensing intervals. Future work to assess the impact of 6MMD on visit burden and resource use would offer a comprehensive view of the benefits to both ART clients and the health system.

## INTRODUCTION

Sustained retention in HIV care is essential to managing the HIV epidemic, particularly in high-burden countries. For individuals on antiretroviral therapy (ART), continuous engagement in care is essential to maintaining viral suppression (1–3), reducing mortality (4), and preventing transmission (5). Frequent clinic visits, however, place a heavy burden on HIV clients (6–9) and health care systems (10) and may hamper retention. To address these challenges, in 2016 the World Health Organization (WHO) recommended differentiated service delivery (DSD) models, including separating clinical consultations from ART dispensing events for stable clients (11). These guidelines advocated extending ART refill intervals to every 3–6 months to ease health system strain and reduce client burden. WHO reaffirmed this guidance in its 2021 guideline revision (12, 13).

Several African countries have now transitioned from three-month dispensing (3MMD) to six-month dispensing (6MMD) (14–16) to reduce structural barriers to care, clinic congestion, and improve efficiency (17–20). In Malawi, 6MMD was introduced nationally in 2019 for stable clients as part of the DSD strategy (21). Initial eligibility criteria included being on ART for at least six months, receiving a first-line ART regimen, having no side effects or opportunistic infections, maintaining viral suppression, adhering to visit schedules, and being at least 24 years old (due to adherence and retention concerns in younger adults) (22, 23). In April 2020, eligibility was expanded in response to the COVID-19 pandemic to clients on dolutegravir-based regimens who weighed at least 20 kg and had been on ART for at least three months, regardless of viral suppression or previous adherence challenges (24). Pregnant and breastfeeding women and children were excluded, remaining on 3MMD. These policy changes were included in the updated 2021 treatment guidance and led to clinic decongestion during the pandemic (25, 26).

Previous studies have shown that 6MMD maintains non-inferior or slightly improved clinical outcomes (viral suppression and retention in care) compared to monthly or three-monthly dispensing (15–18, 20, 27, 28) while reducing patient and health system costs (18). There is limited evidence on its impact on retention using large-scale real-world clinical data, however. In this study, we applied a target trial emulation (TTE) framework to routine observational data from Malawi’s national HIV program to estimate the impact of 6MMD on retention in care compared to shorter dispensing intervals. We also identified client characteristics associated with higher retention to inform implementation strategies and optimize ART delivery.

## METHODS

### Target trial emulation (TTE) and study setting

We used a target trial emulation approach to assess the effect of 6MMD of antiretroviral therapy (ART) on retention among adults with HIV in Malawi (29–33). Target trial emulation (TTE) proposes a hypothetical randomized trial that the investigators would do if they could. The investigators use observational data to mimic that trial as best they can, with a focus on reducing bias through clearly specified eligibility criteria, zero-time, specification of a causal contrast and interventions to be compared, approaches to achieving exchangeability, and clearly defined outcomes.

We applied a TTE analytical approach to compare the effect of 6MMD to standard of care (dispensing intervals of one, two, or three months) on patient retention in care at 12 and 24 months. Table 1 shows the specific components of the target trial and how each was adapted to align with our study using observational data.

**Table 1.** Comparison of target trial protocol and emulated trial for a study of the effect of 6MMD on retention in HIV care at 12 and 24 months in Malawi.

| **Protocol component** | **Target trial protocol** | **Emulated trial** |
| --- | --- | --- |
| **Trial eligibility criteria** | HIV clients $\geq 18$ years of age on ART for at least 6 months who are stable (i.e., virally suppressed) and 6MMD-naïve. | Same as for the trial with the exception that eligibility for 6MMD evidenced by prior 3MMD |
| **Treatment strategy** | Eligible individuals randomized to 6MMD (intervention arm), or non-6MMD [standard 1MMD or 3MMD] (comparison arm) at baseline. | Eligible individuals assigned to: i) 6MMD (intervention arm) if they have evidence of 6-month dispensing experience at eligibility; ii) non-6MMD (comparison arm) if they do not have evidence of 6-month dispensing experience at eligibility. |
| **Treatment assignment** | Eligible individuals randomly assigned to treatment (6MMD) or standard of care (SOC) (non-6MMD) groups. | Randomization was emulated in the observational variant through adjustment for baseline covariates (age, sex, facility type, region, WHO clinical staging, and duration on ART) to address confounding. |
| **Follow-up** | Start: Follow-up begins once an individual is 6MMD-eligible and randomized to an intervention.<br>End: Follow-up ends at death, loss to follow-up, or 36 months, whichever occurs first in the respective trial periods. | Start: Follow up begins at first 6MMD visit in the trial eligibility period for the intervention arm and the first visit in the trial eligibility period for the SOC group.<br>End: Same as the trial, with administrative censoring at 36 months to ensure sufficient follow-up time for outcome assessment. |
| **Outcomes** | Primary outcome: Retention in care at 12 months defined as having a clinic visit between 12-24 after intervention assignment.<br>Secondary outcome: Retention in care at 24 months defined as having a clinic visit between 24-36 after intervention assignment. | Same as target trial protocol. |
| **Causal contrast** | Intention-to-treat (ITT) effect: fixed group assignment based on randomization arm. Treatment effects are evaluated based on original assignment. | Observational equivalent of ITT: The effect of being assigned to 6MMD vs. non-6MMD group at baseline, regardless of subsequent changes in dispensing patterns in the follow-up period. |
| **Statistical analysis** | Risk differences with 95% confidence interval would be estimated. | Poisson regression model with a link identity function and robust variances adjusting for covariates to estimate adjusted risk differences with 95% confidence intervals by pooling together 4 trials each defined by a 6-month eligibility period. |

### Data source

To emulate the trial, we used data from Malawi’s Electronic Medical Record (EMR) system (34, 35) for 26 of the country’s 28 districts between January 2020 and December 2021, which overlapped with 6MMD scaleup. The Neno and Likoma districts were not included in the EMR dataset since the system is implemented only in higher-volume sites, and these districts have relatively low patient volumes.

### Population and eligibility criteria

In our emulated trials, eligibility for 6MMD was defined by the following criteria: being a clinically stable, non-pregnant adult living with HIV (≥18 years) who had been on ART for at least six months, and was 6MMD naïve (no prior exposure to 6MMD) at the time of trial eligibility. In the absence of a viral load test result to assess clinical stability, we assumed individuals were clinically stable if there was evidence of 3MMD in the preceding 6 months at the beginning of each trial period, indicating their eligibility for transition to 6MMD.

The intervention group consisted of participants receiving 6MMD, defined as a six-month ART supply at each dispensing event. The comparison group included participants receiving shorter dispensing intervals (1–3 months).

### Defining the trials and zero time

A target trial has a clearly defined time zero, the point at which individuals become eligible for the trial and are randomized. Immortal time bias, which occurs when follow-up includes periods during which a person cannot experience the outcome. For example, consider the scenario where a patient is eligible for 6MMD at the start of the follow-up period, but only initiates 6MMD two years later. They must have remained in care during this period but could not have experienced the outcome of loss-to-follow-up in that time in our emulation because the outcome we are measuring is retention in care. Anyone who eventually starts 6MMD must, by definition, have remained in care up to that point. This is true even though in real life they are at risk of being lost during that time. This creates “immortal time” where the outcome is artificially improved in the 6MMD group. Because we had no clear time zero for individuals in the emulation, to minimize this bias, we defined four emulated trials using six-calendar-month intervals from January 2020 to December 2021 (Table 1 and S1 Fig), reassessing eligibility to initiate 6MMD at the start of each period to (30, 33, 36), aligning time zero with eligibility.

Within each six-month trial period, eligible individuals were assigned to the 6MMD (intervention) if they received a six-month supply of ART or non-6MMD (comparison) group at time zero (baseline) if they received shorter dispensing intervals (less than 6 months) and followed to assess retention in care. This meant that those in the comparison group had a chance of being included in multiple trials, since anyone who had not already begun 6MMD in an earlier trial was eligible for the next trial. Since experience on 3MMD was used to proxy eligibility for 6MMD, we assessed 6MMD eligibility based on 3MMD experience in the six months preceding each trial. Individuals who initiated ART before or after 1st June 2019 were included and assessed for eligibility in each trial period, provided that they had a clinic visit after this date. This allowed us to check for 3MMD experience in the prior six months to determine 6MMD eligibility for the first trial period (January– June 2020). Baseline was defined as the date the first 6MMD was dispensed for the intervention group and the first clinic visit for the comparison group. An intention-to-treat approach was used, which meant that individuals remained in the group they were assigned at baseline regardless of subsequent switching. Individuals could contribute to multiple trial periods but were only eligible for inclusion in the 6MMD arm once (Fig 1). Once an individual had been included in a 6MMD trial arm, they no longer qualified for subsequent trials as they were no longer naïve to 6MMD.

**Fig 1.**
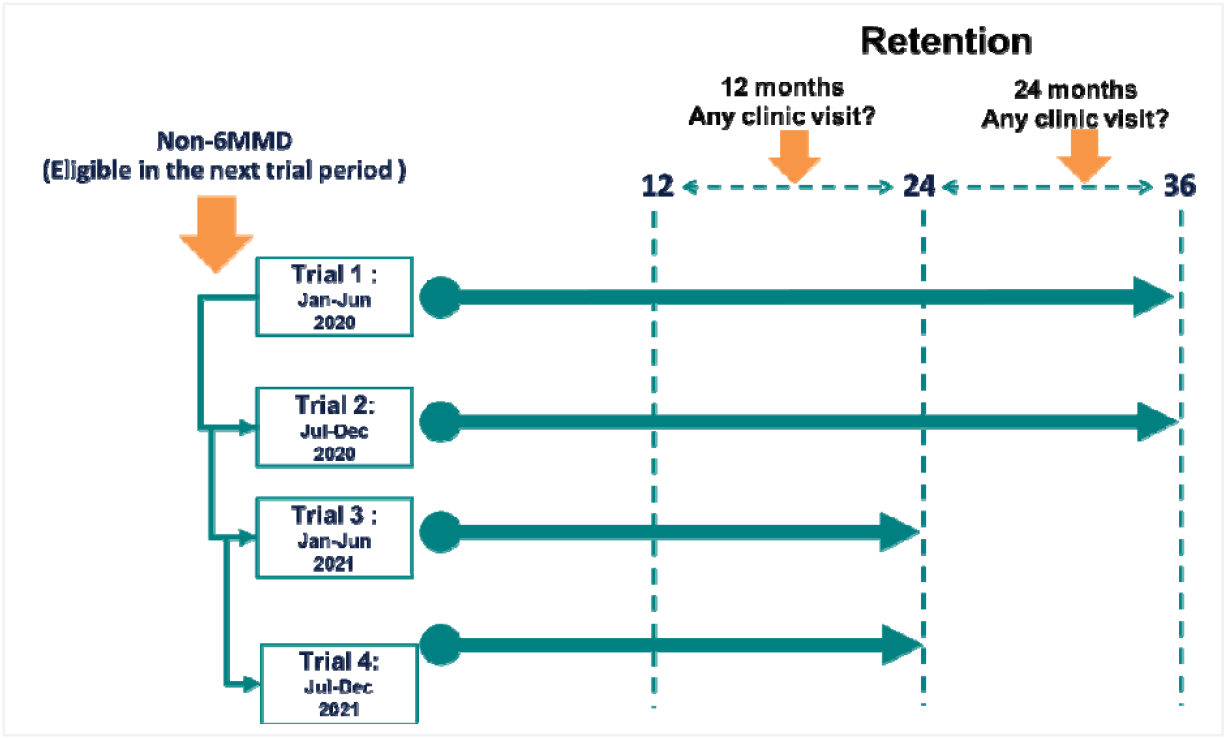
Retention assessment for each emulated trial. **Note:** Retention was assessed for each trial at 12 and 24 months after time zero. The unit of measure is trial-client. Retention at 12 months was defined as having a clinic visit within 12–24 months for trials 1–4, and retention at 24 months was defined as having a clinic visit within 24–36 months for trials 1–2.

Individuals were followed up for up to 24 months with retention at 12 months defined by a recorded clinic visit between 12-24 months and for Trials 1 and 2, follow-up was extended to 36 months to allow sufficient follow-up time to assess retention at 24 months defined by a recorded clinic visit between 24-36 months (see outcome measurement in Fig 1)..

### Baseline characteristics and outcome measurement

Baseline characteristics were described for all patients and measured at baseline (defined above) of the first eligible trial enrollment. Characteristics of patients included age group at first eligible date, sex, WHO HIV clinical stage at ART initiation, duration on ART at first eligible trial enrollment, facility type (primary healthcare (PHC) facilities versus hospitals), and region (north, central and south regions).

The primary outcome was retention in care, defined as having a clinic visit within 12–24 months for 12-month retention or 24-36 months for 24-month retention after enrollment into each respective trial (S1 Table). Retention at 12 months was assessed across all four trial periods, while retention at 24 months was limited to the first two trial periods.

### Statistical analysis

We first summarised baseline characteristics of patients in both 6MMD and non-6MMD groups using median values with interquartile ranges (IQR) for continuous variables and frequencies with percentages for categorical variables. Since patients could be eligible for multiple trials, our unit of measure in the analysis was a trial-client as opposed to a unique participant. After trial-clients for all four trials were assigned, we estimated crude (RD) and adjusted risk differences (aRD), and corresponding 95% confidence intervals for retention at 12 (trial 1-4) and 24 (trial 1-2) months.

Estimates were obtained using a generalized Poisson regression model with an identity link function in each trial period, comparing 6MMD and non-6MMD groups. We adjusted for relevant demographic (age, sex, clinical (WHO clinical stage at ART initiation, duration on ART), and facility covariates (facility type and regional location), considered as potential confounding individual and facility factors.

We combined the data across all trials to estimate a pooled adjusted risk difference, accounting for within-subject variation across trials using robust variances. We conducted stratified analysis by age and sex to determine whether the effect of 6MMD varied across these demographic groups. For the causal contrasts, we emulated the intention-to-treat estimate, where treatment effects are evaluated based on original randomization group. The observational equivalent of the intention-to-treat effect was estimated by analysing participants based on the original treatment assignment for each emulated trial, regardless of any later changes in dispensing frequency.

### Ethical considerations

Permission to access anonymized routine data from the EMR system was obtained from the Malawi Ministry of Health (MOH). The study received ethical approval from three institutional review boards: the Malawi National Health Sciences Research Committee (NHSRC Ref: 2672), Boston University Institutional Review Board (IRB Number: H-38822), and the University of the Witwatersrand Human Research Ethics Committee (Ref: M190451). Only anonymised data was accessed by the research team and analysis outputs for this were extracted from the server.

## RESULTS

### Study population

The exported dataset included clinic visit history data for 735,694 patients who had a clinic visit between January 2020 and December 2021 and were assessed for eligibility for at least one trial. The dataset covered 239 healthcare facilities across 26 of the 28 districts in Malawi, covering the Northern, Central, and Southern regions.

Since patients could be included in more than one trial, the unit of measure was a “trial-client” rather than a unique patient. In total, 2,431,316 trial-clients were assessed for eligibility during the trial enrollment periods (Fig2, S1-S2 Figure). We excluded those with missing facility start date in the EMR (n = 4,414; 0.2%), or with ART initiation occurring before EMR start date (n = 1,271,724; 52%), leaving a total of 1,155,178 trial-clients (47.5%). We then excluded those not eligible for 6MMD, including clients on ART for <6 months (n = 163,404; 6.7%), younger than 18 years (n = 83,744; 3.4%), pregnant at baseline (n = 33,104; 1.4%), with prior exposure to 6MMD rollout (n = 525,834; 21.6%), or with no prior 3MMD exposure (n = 84,926; 3.5%).

**Fig 2.**
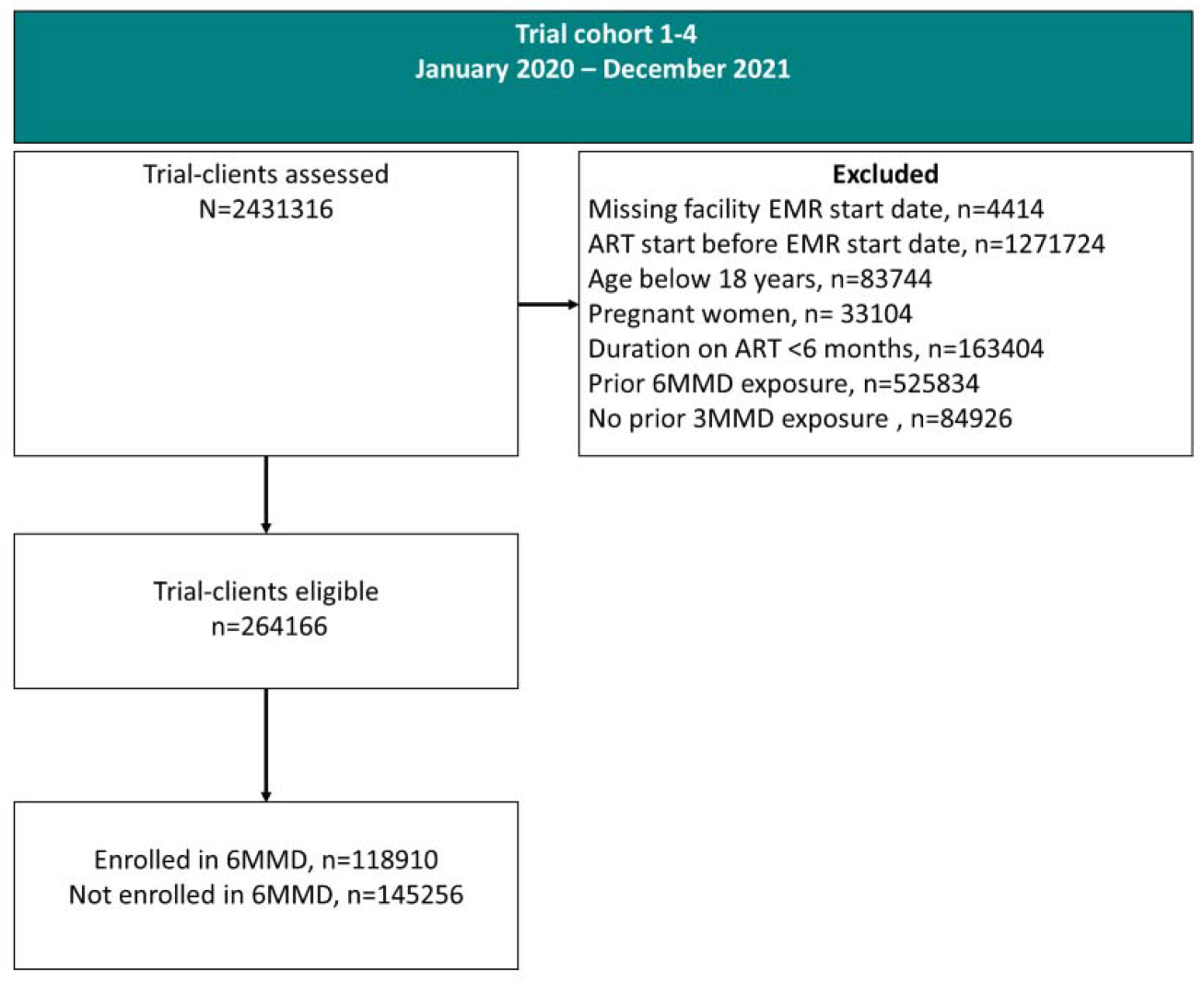
Consolidated flow diagram of trial-client eligibility for all four target trials. Note: The consort diagram shows all trial-clients assessed at baseline in all trial periods. Remaining eligible trial-clients after applying the exclusions were included in follow-up and allocated to intervention or control arms.

After all exclusions, a total of 264,166 trial-clients (10.9% of all assessed), corresponding to 159,801 unique patients, were eligible for inclusion. Among these, 118,910 (45.0%) were enrolled in 6MMD, while 145,256 (55.0%) were not enrolled.

### Baseline characteristics

Among the 159,801 patients eligible for 6MMD who were included in this analysis, 74% (n=118,910) ever experienced 6MMD and 26% (n=40,891) never experienced 6MMD (Table 2). Most participants were female (60.7%), and had a median age of 37 years (IQR: 30-44). Nearly 54% accessed care at primary healthcare clinics, and the majority (59.3%) were from facilities located in the southern region of the country. Those ever on 6MMD were older, with a median age of 38 years (IQR: 31–45) compared to 33 years (IQR: 26–40) among those never on 6MMD. Younger patients (aged 17–24 years) represented more than half the non-6MMD group (56.2%) compared to just over a third of the 6MMD group (36.3%); the 6MMD group, in contrast, included more patients aged 50 years and above (8.9% vs 4.5%). Those ever on 6MMD had been on ART longer (median, IQR: 37 months, 17– 67) compared to those never on 6MMD (24 months, 12–51). While most participants in both groups were either asymptomatic or had mild symptoms (WHO Stage 1 or 2) at ART initiation, there was a slightly higher proportion of advanced disease among those ever on 6MMD (19.1%) compared to those not on 6MMD (13.6%). The cohort characteristics based on trial-clients also showed similar distribution across all variables (S2 Table).

**Table 2.**
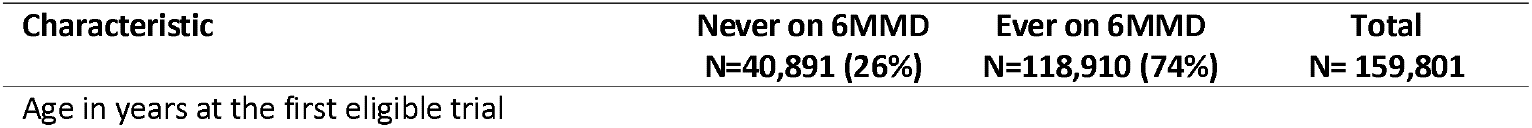
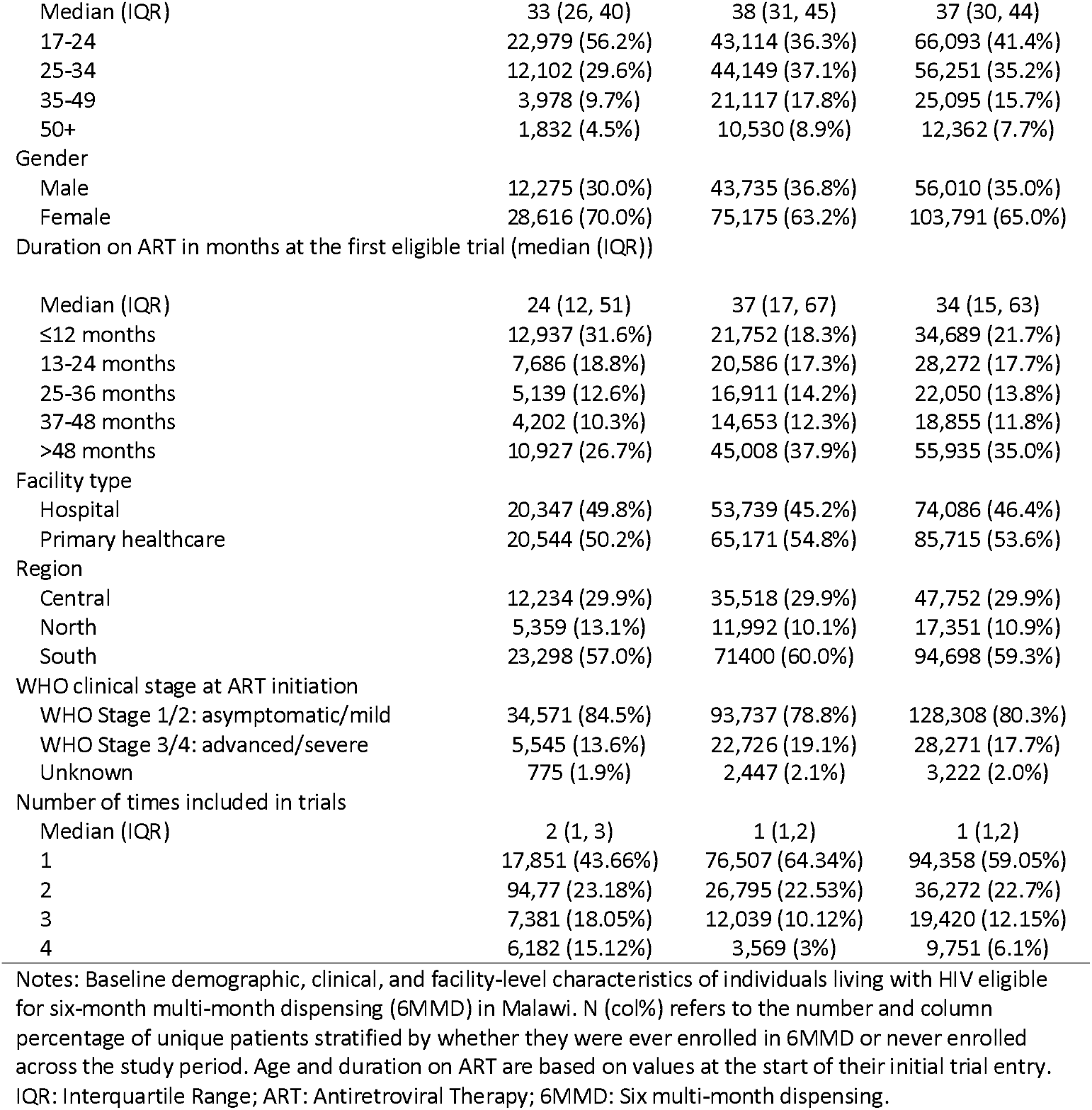
Characteristics of unique patients eligible for 6MMD a target trial emulation study of the effect of 6 monthly dispensing of HIV treatment in Malawi.

| Characteristic | Never on 6MMD<br>N=40,891 (26%) | Ever on 6MMD<br>N=118,910 (74%) | Total<br>N= 159,801 |
| --- | --- | --- | --- |
| Age in years at the first eligible trial |  |  |  |
| Median (IQR) | 33 (26, 40) | 38 (31, 45) | 37 (30, 44) |
| 17-24 | 22,979 (56.2%) | 43,114 (36.3%) | 66,093 (41.4%) |
| 25-34 | 12,102 (29.6%) | 44,149 (37.1%) | 56,251 (35.2%) |
| 35-49 | 3,978 (9.7%) | 21,117 (17.8%) | 25,095 (15.7%) |
| 50+ | 1,832 (4.5%) | 10,530 (8.9%) | 12,362 (7.7%) |
| Gender |  |  |  |
| Male | 12,275 (30.0%) | 43,735 (36.8%) | 56,010 (35.0%) |
| Female | 28,616 (70.0%) | 75,175 (63.2%) | 103,791 (65.0%) |
| Duration on ART in months at the first eligible trial (median (IQR)) |  |  |  |
| Median (IQR) | 24 (12, 51) | 37 (17, 67) | 34 (15, 63) |
| ≤12 months | 12,937 (31.6%) | 21,752 (18.3%) | 34,689 (21.7%) |
| 13-24 months | 7,686 (18.8%) | 20,586 (17.3%) | 28,272 (17.7%) |
| 25-36 months | 5,139 (12.6%) | 16,911 (14.2%) | 22,050 (13.8%) |
| 37-48 months | 4,202 (10.3%) | 14,653 (12.3%) | 18,855 (11.8%) |
| >48 months | 10,927 (26.7%) | 45,008 (37.9%) | 55,935 (35.0%) |
| Facility type |  |  |  |
| Hospital | 20,347 (49.8%) | 53,739 (45.2%) | 74,086 (46.4%) |
| Primary healthcare | 20,544 (50.2%) | 65,171 (54.8%) | 85,715 (53.6%) |
| Region |  |  |  |
| Central | 12,234 (29.9%) | 35,518 (29.9%) | 47,752 (29.9%) |
| North | 5,359 (13.1%) | 11,992 (10.1%) | 17,351 (10.9%) |
| South | 23,298 (57.0%) | 71400 (60.0%) | 94,698 (59.3%) |
| WHO clinical stage at ART initiation |  |  |  |
| WHO Stage 1/2: asymptomatic/mild | 34,571 (84.5%) | 93,737 (78.8%) | 128,308 (80.3%) |
| WHO Stage 3/4: advanced/severe | 5,545 (13.6%) | 22,726 (19.1%) | 28,271 (17.7%) |
| Unknown | 775 (1.9%) | 2,447 (2.1%) | 3,222 (2.0%) |
| Number of times included in trials |  |  |  |
| Median (IQR) | 2 (1, 3) | 1 (1,2) | 1 (1,2) |
| 1 | 17,851 (43.66%) | 76,507 (64.34%) | 94,358 (59.05%) |
| 2 | 94,77 (23.18%) | 26,795 (22.53%) | 36,272 (22.7%) |
| 3 | 7,381 (18.05%) | 12,039 (10.12%) | 19,420 (12.15%) |
| 4 | 6,182 (15.12%) | 3,569 (3%) | 9,751 (6.1%) |

Notes: Baseline demographic, clinical, and facility-level characteristics of individuals living with HIV eligible for six-month multi-month dispensing (6MMD) in Malawi. N (col%) refers to the number and column percentage of unique patients stratified by whether they were ever enrolled in 6MMD or never enrolled across the study period. Age and duration on ART are based on values at the start of their initial trial entry. IQR: Interquartile Range; ART: Antiretroviral Therapy; 6MMD: Six multi-month dispensing.

### Retention in care at 12 and 24 months

The pooled mean proportion of individuals retained in care (Fig 3a) at 12 months for the 6MMD group (90.9%) was 3.5% higher than for the non-6MMD group (87.3%). Similarly, at 24 months (Fig 3b) the 6MMD group had a slightly higher mean proportion of retention of 84.3% than did the non-6MMD group (80.8%). This finding was similar across all the trials at both the 12- and 24-month outcome assessment.

**Fig 3.**
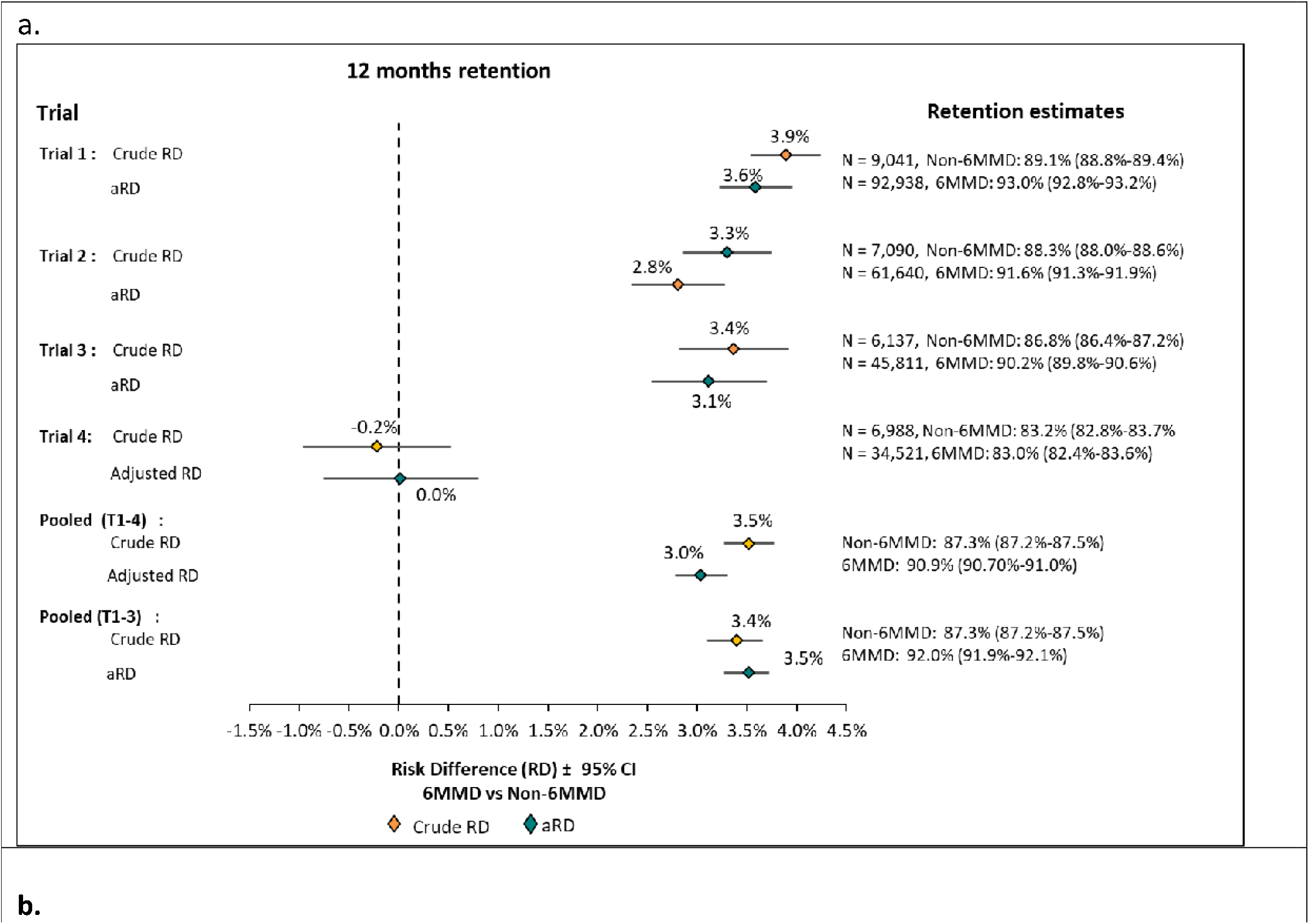
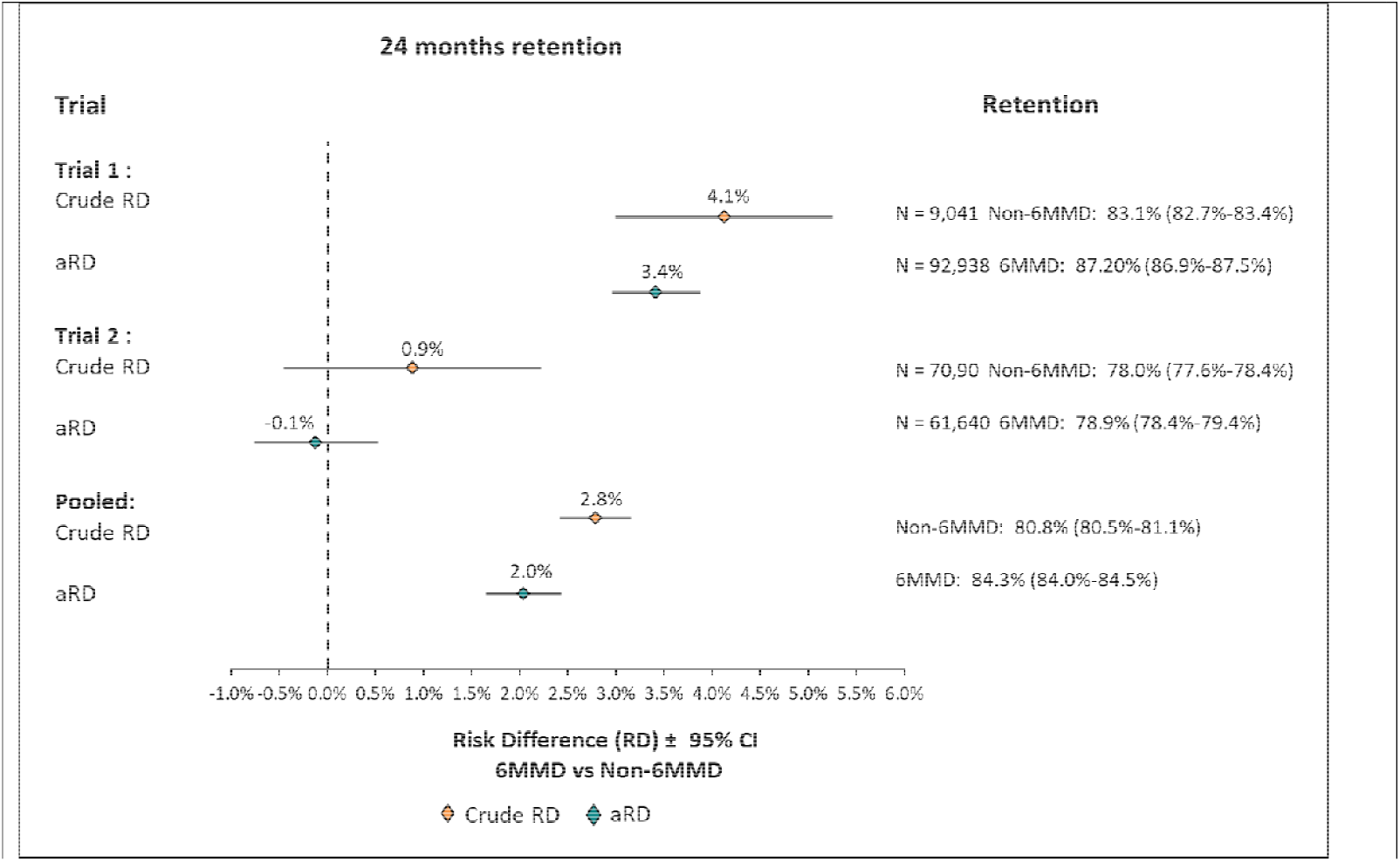
Individual and pooled crude RD and aRD of 12 and 24 month retention in all the trials. Notes: Forest plots of individual trial estimates and pooled estimates of the risk difference in retention in care at 12 and 24 months. Each horizontal line represents one trial, crude (RD)and adjusted risfk differences (aRD) and 95% confidence intervals (CI). The green diamond represents theadjusted RD and the orage one is for the crude RD. Positive RDs indicate higher retention in those with 6MMD compared with those non-6MMD group.

Differences in retention estimates

The adjusted risk difference (aRD) for 12-month retention among patients ever on 6MMD (Fig 3a and Table 3) compared to those never on 6MMD pooled across the four trials was 3.0% (95% CI: 2.8%–3.3%). For 24-month retention, the adjusted RD was slightly lower at 2.0% (95% CI: 1.7%– 2.4%) (Fig 3b and Table 3). These estimates were adjusted for age at first eligible trial, sex, WHO stage at ART initiation, duration on ART at first eligible trial, facility type, and the country’s regions. Patient trial estimates showed similar retention estimates and risk differences (S3 Table and S4 Table). Because trial 4 showed no significant differences between the two arms compared with the other trials, we pooled trials 1–3 to reduce heterogeneity, estimating retention (92%; 95% CI: 91.9-92.1) and the adjusted RD (3.5%, 95% CI: 3.3-3.8).

**Table 3.** Pooled adjusted risk differences of 12- and 24-month retention across cohort characteristics in Malawi.

| Characteristic | 12 months |  |  |  | 24 months |  |  |  |
| --- | --- | --- | --- | --- | --- | --- | --- | --- |
|  | Not Retained | Retained | Total | aRD [95% CI] | Not Retained | Retained | Total | aRD [95% CI] |
| **N (trial-clients)** | 29,256 | 234,910 | 264,166 |  | 29,923 | 140,786 | 170,709 |  |
| **Dispensing interval** |  |  |  |  |  |  |  |  |
| Non-6MMD | 16,734 (20.0%) | 66,942 (80.0%) | 83,676 | Reference | 11,922 (14.2%) | 23,745 (28.4%) | 35,667 | Reference |
| 6MMD | 12,522 (6.9%) | 167,968 (93.1%) | 180,490 | 3.0% (2.8%-3.3%) | 18,001 (10.0%) | 117,041 (64.8%) | 135,042 | 2.0% (1.7%-2.4%) |
| **Age group at first eligible trial** |  |  |  |  |  |  |  |  |
| 18–24 | 15,298 (13.3%) | 99,970 (86.7%) | 115,268 | Reference | 14,234 (12.3%) | 52,891 (45.9%) | 67,125 | Reference |
| 25–34 | 9,087 (9.9%) | 82,655 (90.1%) | 91,742 | 3.4% (3.0%-3.7%) | 9,875 (10.8%) | 51,910 (56.6%) | 61,785 | 4.8% (4.3%-5.3%) |
| 35–49 | 3,153 (8.2%) | 35,171 (91.8%) | 38,324 | 5.4% (5.0%-5.8%) | 3,749 (9.8%) | 24,060 (62.8%) | 27,809 | 7.5% (6.9%-8.2%) |
| 50+ | 1,718 (9.1%) | 17,114 (90.9%) | 18,832 | 4.4% (3.8%-5.0%) | 2,065 (11.0%) | 11,925 (63.3%) | 13,990 | 6.1% (5.3%-6.9%) |
| **Sex** |  |  |  |  |  |  |  |  |
| Male | 11,576 (13.5%) | 74,266 (86.5%) | 85,842 | Reference | 11,441 (13.3%) | 45,910 (53.5%) | 57,351 | Reference |
| Female | 17,680 (9.9%) | 160,644 (90.1%) | 178,324 | 4.7% (4.4%-5.1%) | 18,482 (10.4%) | 94,876 (53.2%) | 113,358 | 5.1% (4.7%-5.6%) |
| **Duration on ART at first eligible trial in months** |  |  |  |  |  |  |  |  |
| ≤12 | 7,714 (14.7%) | 44,699 (85.3%) | 52,413 | Reference | 4,892 (9.3%) | 16,102 (30.7%) | 20,994 | Reference |
| 13–24 | 6,321 (13.4%) | 40,686 (86.6%) | 47,007 | 1.1% (0.6%-1.6%) | 6,476 (13.8%) | 23,796 (50.6%) | 30,272 | 0.7% (-0.2%-1.5%) |
| 25–36 | 4,311 (11.6%) | 32,770 (88.4%) | 37,081 | 2.8% (2.3%-3.4%) | 5,095 (13.7%) | 21,542 (58.1%) | 26,637 | 2.6% (1.7%-3.4%) |
| 37–48 | 3,294 (10.2%) | 28,874 (89.8%) | 32,168 | 4.1% (3.6%-4.6%) | 3,879 (12.1%) | 19,294 (60.0%) | 23,173 | 4.8% (4.0%-5.7%) |
| >48 | 7,616 (8.0%) | 87,881 (92.0%) | 95,497 | 6.0% (5.6%-6.4%) | 9,581 (10.0%) | 60,052 (62.9%) | 69,633 | 7.7% (6.9%-8.4%) |
| **Facility type** |  |  |  |  |  |  |  |  |
| Hospital | 14,742 (11.4%) | 114,169 (88.6%) | 128,911 | Reference | 15,123 (11.7%) | 67,392 (52.3%) | 88,194 | Reference |
| Primary healthcare | 14,514 (10.7%) | 120,741 (89.3%) | 135,255 | 1.4% (1.1%-1.7%) | 14,800 (10.9%) | 73,394 (54.3%) | 82,515 | 2.6% (2.1%-3.0%) |
| **Region** |  |  |  |  |  |  |  |  |
| Central | 9,860 (12.2%) | 71,284 (87.8%) | 81,144 | Reference | 10,757 (13.3%) | 41,660 (51.3%) | 52,417 | Reference |
| North | 3,810 (13.2%) | 25,107 (86.8%) | 28,917 | -0.4% (-1.0%-0.1%) | 3,561 (12.3%) | 15,055 (52.1%) | 18,616 | 2.6% (1.8%-3.4%) |
| South | 15,586 (10.1%) | 138,519 (89.9%) | 154,105 | 3.2% (2.8%-3.5%) | 15,605 (10.1%) | 84,071 (54.6%) | 99,676 | 6.7% (6.2%-7.2%) |
| **WHO clinical stage at ART initiation** |  |  |  |  |  |  |  |  |
| Stage I/II | 24,358 (11.5%) | 187,450 (88.5%) | 211,808 | Reference | 24,218 (11.4%) | 109,348 (51.6%) | 133,566 | Reference |
| Stage III/IV | 4,558 (9.7%) | 42,316 (90.3%) | 46,874 | -0.2% (-0.5%-0.2%) | 5,371 (11.5%) | 28,534 (60.9%) | 33,905 | -0.5% (-1.0%-0.1%) |
| Missing | 340 (6.2%) | 5,144 (93.8%) | 5,484 |  | 334 (6.1%) | 2,904 (53.0%) | 3,238 |  |

Notes: Values are presented as n (%) retained at 12 and 24 months for each characteristic, with the unit of measure being the trial-client. Adjusted Risk differences (aRD) compare retention between each category and the designated reference group, adjusting for including ever on 6MMD, age at first eligible trial, sex, duration on ART at first eligible trial, facility type, and region.

### Stratified analyses

Retention at 12 months was consistently higher among patients ever on 6MMD compared to those never on 6MMD across all age groups at first eligible trial (Table 5). Retention was 87.6% vs. 86.5% among those aged 18–34 years (aRD 1.6%, 95% CI: 1.1–2.1); 92.0% vs. 89.2% among those aged 35– 44 years (3.2%, 2.8–3.7); 93.9% vs. 90.4% among those aged 45–54 years (3.4%, 2.8–4.1); and 93.0% vs. 89.5% among those aged 55+ years (3.3%, 2.4–4.2). At 24 months, similar patterns were observed. By sex, retention was also higher among those ever on 6MMD for both males and females. At 12 months, retention was 89.9% vs. 84.7% (4.9%, 4.4–5.4) among males, while 91.7% vs. 89.5% (1.3%, 1.0–1.6) among females. At 24 months, retention remained higher among those on 6MMD: 84.0% vs. 77.0% among males (6.5%, 5.8–7.3) and 86.3% vs. 82.4% among females (2.3%, 1.8–2.8).

**Table 5.**
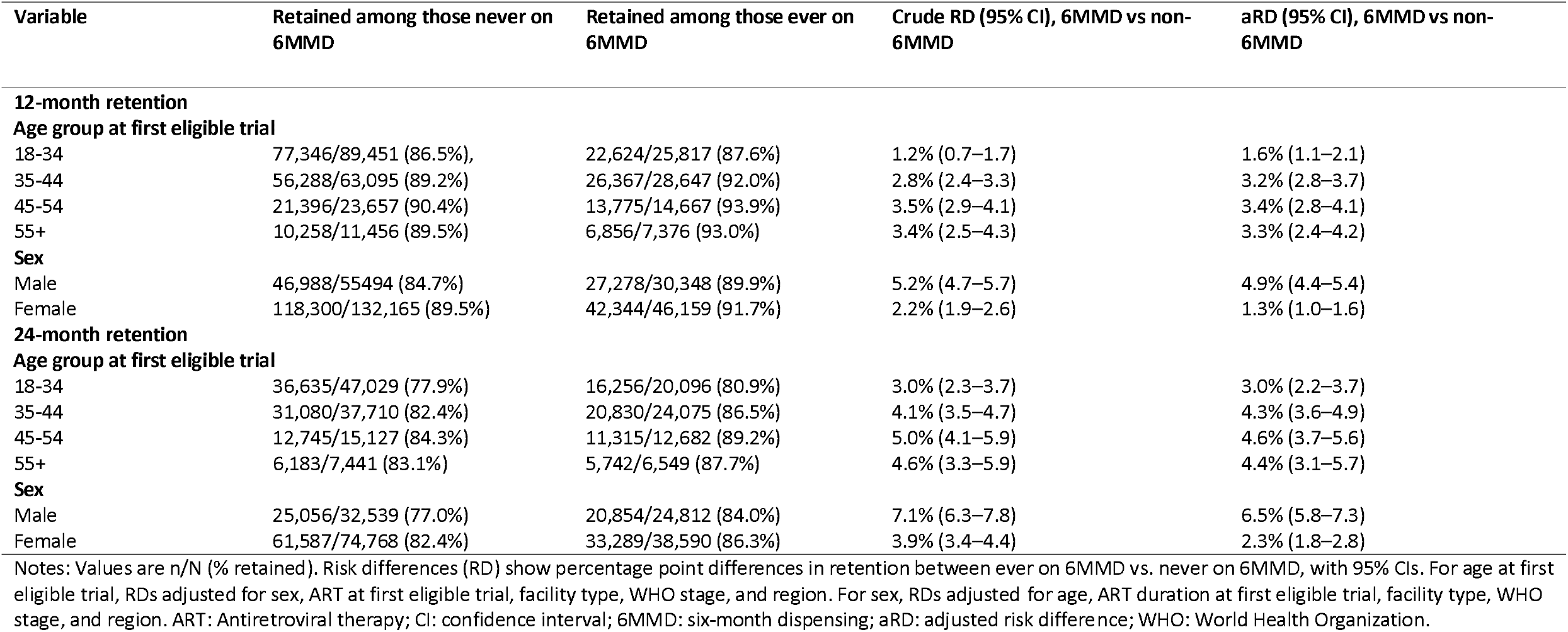
Retention at 12- and 24-months comparing patients ever on 6MMD to those never on 6MMD, stratified by age and sex.

| Variable | Retained among those never on 6MMD | Retained among those ever on 6MMD | Crude RD (95% CI), 6MMD vs non-6MMD | aRD (95% CI), 6MMD vs non-6MMD |
| --- | --- | --- | --- | --- |
| **12-month retention** |  |  |  |  |
| **Age group at first eligible trial** |  |  |  |  |
| 18-34 | 77,346/89,451 (86.5%), | 22,624/25,817 (87.6%) | 1.2% (0.7–1.7) | 1.6% (1.1–2.1) |
| 35-44 | 56,288/63,095 (89.2%) | 26,367/28,647 (92.0%) | 2.8% (2.4–3.3) | 3.2% (2.8–3.7) |
| 45-54 | 21,396/23,657 (90.4%) | 13,775/14,667 (93.9%) | 3.5% (2.9–4.1) | 3.4% (2.8–4.1) |
| 55+ | 10,258/11,456 (89.5%) | 6,856/7,376 (93.0%) | 3.4% (2.5–4.3) | 3.3% (2.4–4.2) |
| **Sex** |  |  |  |  |
| Male | 46,988/55,494 (84.7%) | 27,278/30,348 (89.9%) | 5.2% (4.7–5.7) | 4.9% (4.4–5.4) |
| Female | 118,300/132,165 (89.5%) | 42,344/46,159 (91.7%) | 2.2% (1.9–2.6) | 1.3% (1.0–1.6) |
| **24-month retention** |  |  |  |  |
| **Age group at first eligible trial** |  |  |  |  |
| 18-34 | 36,635/47,029 (77.9%) | 16,256/20,096 (80.9%) | 3.0% (2.3–3.7) | 3.0% (2.2–3.7) |
| 35-44 | 31,080/37,710 (82.4%) | 20,830/24,075 (86.5%) | 4.1% (3.5–4.7) | 4.3% (3.6–4.9) |
| 45-54 | 12,745/15,127 (84.3%) | 11,315/12,682 (89.2%) | 5.0% (4.1–5.9) | 4.6% (3.7–5.6) |
| 55+ | 6,183/7,441 (83.1%) | 5,742/6,549 (87.7%) | 4.6% (3.3–5.9) | 4.4% (3.1–5.7) |
| **Sex** |  |  |  |  |
| Male | 25,056/32,539 (77.0%) | 20,854/24,812 (84.0%) | 7.1% (6.3–7.8) | 6.5% (5.8–7.3) |
| Female | 61,587/74,768 (82.4%) | 33,289/38,590 (86.3%) | 3.9% (3.4–4.4) | 2.3% (1.8–2.8) |

Notes: Values are n/N (% retained). Risk differences (RD) show percentage point differences in retention between ever on 6MMD vs. never on 6MMD, with 95% CIs. For age at first eligible trial, RDs adjusted for sex, ART at first eligible trial, facility type, WHO stage, and region. For sex, RDs adjusted for age, ART duration at first eligible trial, facility type, WHO stage, and region. ART: Antiretroviral therapy; CI: confidence interval; 6MMD: six-month dispensing; aRD: adjusted risk difference; WHO: World Health Organization.

## DISCUSSION

This study evaluated the impact of 6MMD on retention in care among patients on ART in Malawi using routine EMR data. Our findings show that patients on 6MMD have equal or slightly higher retention in care at 12 and 24 months than patients on shorter dispensing intervals (1–3 months).

The TTE approach used in this analysis approximated the design of a randomized trial to estimate the causal effect of 6MMD on retention in care using routine EMR data from Malawi. By using the TTE framework (29), we explicitly defined the eligibility, treatment strategies and follow⍰up start times ensuring each person’s time at risk is correctly defined before 6MMD exposure to minimise immortal time bias, a common source of overestimation in cohort studies (30–32). And by structuring four sequential six-month trials and using robust standard errors to account for within-subject correlation, the TTE leveraged all available data to increase precision (30, 32).

Our findings on the benefits of 6MMD reaffirm those reported in prior controlled clinical trials and observational studies that did not minimize immortal time bias. The INTERVAL cluster⍰randomized trial found 6⍰monthly dispensing being non⍰inferior to standard care and suggested improved retention among stable patients in Malawi and Zambia (18). Cluster randomized trials in other sub-Saharan countries also found that retention with 6MMD was non-inferior to 3MMD when delivered through DSD models, including community ART refill groups in Zimbabwe (17), adherence clubs (37) and facility-based care (18) in South Africa, and community pick-up points in Lesotho (20). This has also been reported in other observational studies (15, 16, 27, 28). To our knowledge, however, this is the first study to use large-scale, routine national data from Malawi’s HIV program to evaluate the impact of 6MMD on retention in care using a TTE approach to reduce the biases inherent in observational studies.

Our results support WHO guidelines advocating longer dispensing intervals for stable patients to alleviate clinic congestion and improve access (11–13) and the continued scale-up of differentiated service delivery (DSD) models which incorporate the use of 6MMD to enhance ART retention, particularly in resource-limited settings (38). By extending ART refills to six months, clinically stable patients spend less time and incur fewer costs on travel and waiting, which can promote continuous engagement in care (17–20). For health facilities, fewer routine dispensing visits may free up staff and resources for patients requiring more intensive services (39–42). 6MMD seems to have performed well during the COVID-19 pandemic despite drug stock-outs, inconsistent adherence to eligibility criteria by providers, and missed clinic visits for ART refills and viral load sample collection (43).

We found that 6MMD’s association with retention in care was greater among older female patients. Overall predictors of retention at both 12 and 24 months included 6MMD, older age, female sex long⍰term ART use attendance at primary health clinics, and accessing care in the country’s southern region. Consistent with prior studies in rural Malawi (44), female patients had higher retention than males, likely due to known differences in health⍰seeking behaviour and social support (45). Men’s higher attrition, also reported in other studies(44), suggests that while 6MMD improves treatment access and convenience, it does not necessarily address underlying sex-specific barriers to optimal ART outcomes. Older age groups (35–49 and ≥50 years) also showed greater retention, consistent with findings from studies conducted in Zambia (46) and Zimbabwe (17), potentially because younger patients face barriers such as stigma, mobility, and competing responsibilities (47). Baseline WHO clinical stage at ART initiation did not predict retention in our 6MMD cohort at either 12- or 24 months, in contrast to findings of an earlier study (44) in Malawi’s rural Chiradzulu District, which found that WHO stage 3/4 was independently associated with higher attrition. The higher proportion of advanced HIV disease among those ever on 6MMD, however, suggests the program may be helping patients achieve clinical stability and 6MMD eligibility.

Despite the strength of this study of using a rigorous target trial emulation methodology, some limitations should be noted. First, and potentially most important, the TTE approach cannot fully eliminate bias from unmeasured confounding factors which we could not adjust for. It is likely that clinicians’ judgments as to whether to offer 6MMD to individual patients were based on expectations of future retention and adherence, concentrating higher risk patients in the non-6MMD group. While this limitation may cast doubt on the observed increase in retention for those on 6MMD, it does not negate the core finding that 6MMD is non-inferior to shorter dispensing periods. With equal if not better retention, the other benefits of 6MMD (lower costs, less clinic congestion, etc.) would make it a preferable strategy even if it does not improve clinical outcomes.

Second, missing data in electronic medical records led to substantial exclusions including data prior to EMR facility specific start dates, which may have introduced selection bias. Limited viral load test results in the EMR system also made it impossible to examine the impact of 6MMD on viral suppression as another important treatment outcome at the time of analysis. Third, relying on prior 3⍰month dispensing as a proxy for clinical stability may misclassify 6MMD eligibility, which could potentially lead to biased estimates. Finally, administrative censoring constrained later trials’ follow⍰up, which could attenuate 24⍰month retention estimates and limit generalizability beyond the study period.

## CONCLUSION

In many countries, including Zambia (48), Nigeria (49), and Ethiopia(50), in addition to Malawi (43), a majority of individuals on ART are now receiving 6 months of medications at once, as per government guidelines. Confidence that 6MMD is sustaining HIV treatment outcomes, while also achieving other benefits, is thus of critical importance. Using a target trial emulation approach and national routine EMR data, we found that 6MMD achieved comparable or slightly higher retention rates at 12 and 24 months compared to shorter dispensing intervals in Malawi. By emulating multiple six⍰month trials using routine EMR data, we approximated causal effects in a real⍰world context while enhancing external validity. The large sample size and adjustment for key confounders strengthen the inferences of our findings. These findings reaffirm results from randomized trials and demonstrate the desirability of scaling up 6MMD within national HIV programs.

## Data availability

The de-identified data used in this study were provided by the Ministry of Health’s Department of HIV and AIDS through CHAI. To protect patient privacy, strict confidentiality protocols and ethical approvals were in place. Our access to the data was limited to a secure remote server, where all data processing and analysis were conducted and only the final results were permitted to be extracted.

As such, requests for data access can be directed to the Department of HIV/AIDS at the Ministry of Health (https://hiv.health.gov.mw/). The analytical scripts used for statistical analysis in this study are publicly available at: https://github.com/kshumba/TTE_6MMD_Malawi_2025.

## Authors’ contributions

KS, ANH, IM, LJ and MPF conceptualised the study. IM, ANH, SP and TT facilitated remote data access. The TTE framework was developed by KS, MPF, ANH, LJ, EK and IM. KS conducted the data analysis with guidance from MPF, ANH, IM, LS and EK. The initial draft was written by KS and critically reviewed by MPF, SR, LJ, SP, ANH, EK, and SN. All authors contributed to reviewing early analyses and provided critical feedback on the manuscript.

## Acknowledgements

The authors gratefully acknowledge the Department of HIV and AIDS at the Ministry of Health Malawi for granting access to data extracts and for ongoing technical support. We also thank the CHAI team for their assistance in facilitating accessing and providing support in interpreting analytical outputs. Finally, we extend our appreciation to the clinical staff delivering care to people living with HIV in Malawi’s public health facilities.

