## Supplementary Material for "The Impact of 6-Month ART Dispensing (6MMD) on Retention in Malawi’s HIV Program: A Target Trial Emulation Study"

A. **SUPPORTING INFORMATION**

**Figures:**

**S1 Figure**. **Eligibility assessment at trial baseline**
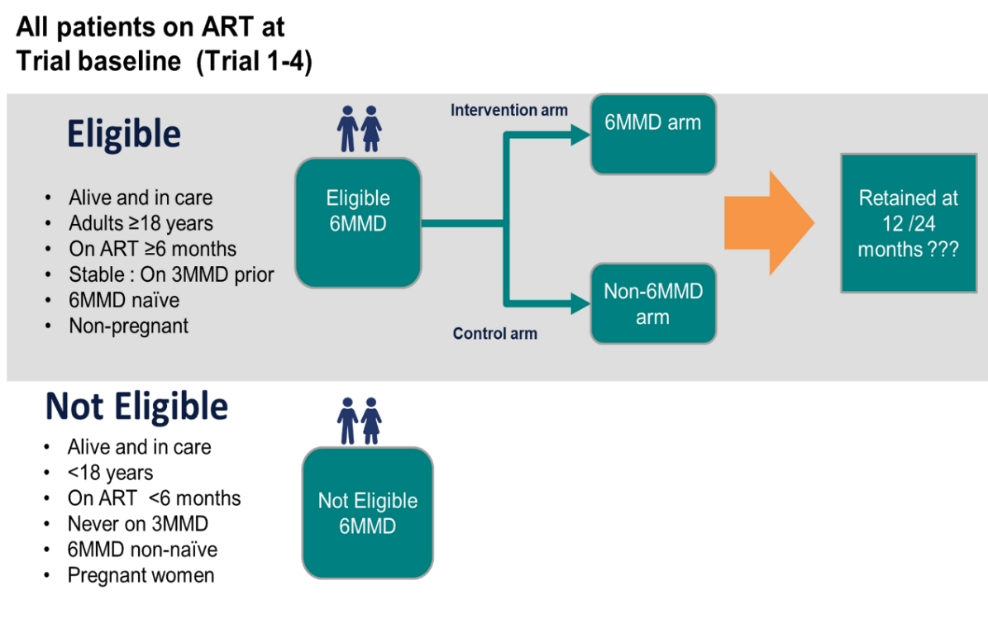

**Note**: The unit of measure is trial-client. Trial-clients were assessed for inclusion at baseline of each trial period. Exclusions were applied. Remaining trial-clients meeting all criteria were eligible for allocation to intervention or control arms.

**S2 Figure. Flow diagrams of trial-client eligibility and follow-up for each target trials**

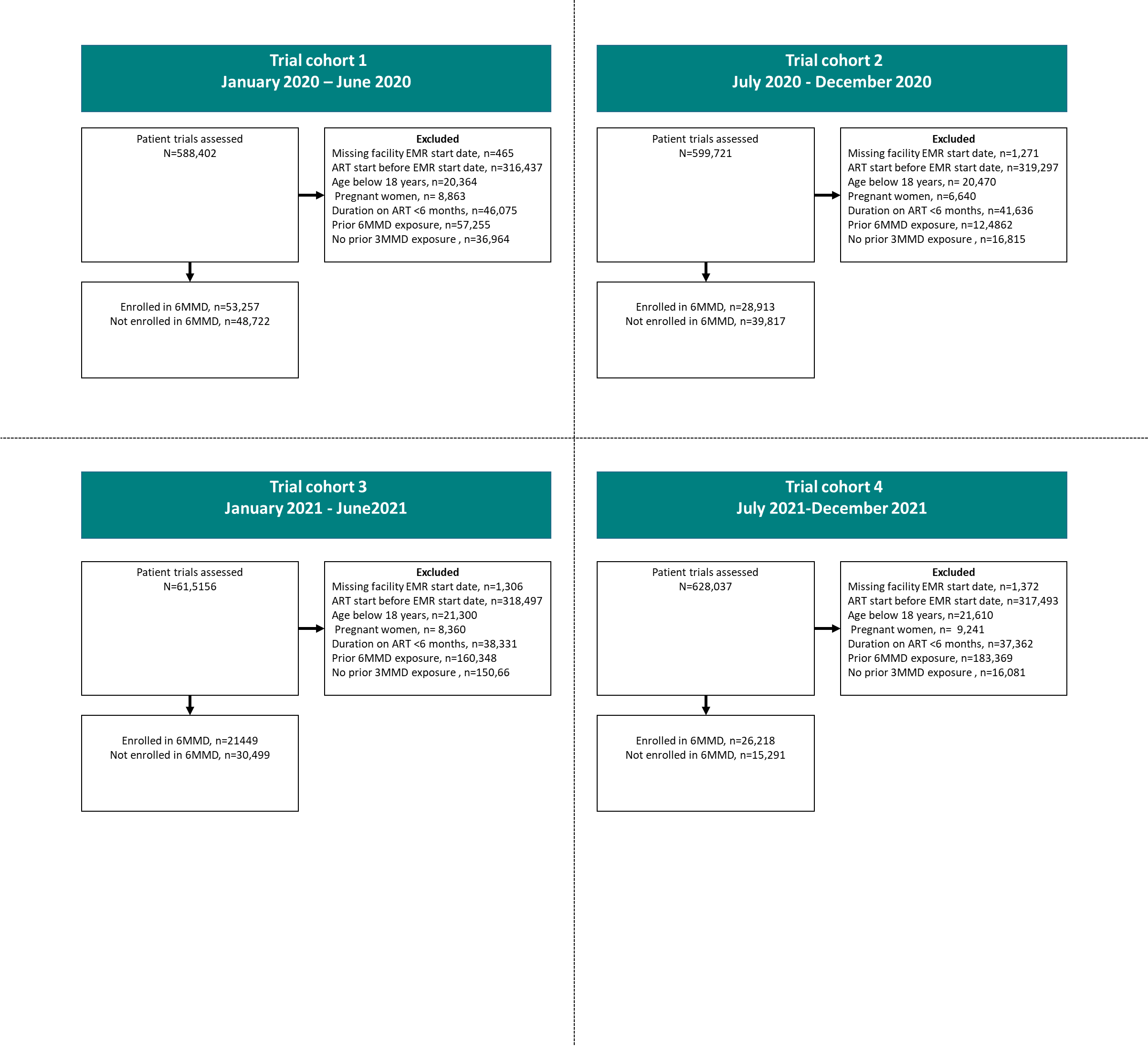

**Notes:** The consort diagrams show all trial-clients assessed at baseline in each trial period. Remaining eligible trial-clients after applying the exclusions were included in follow-up and allocated to intervention or control arms.

B. **TABLES**

**S1 Table. Summary of trial definitions and follow-up criteria**

| **Trial period** | **Baseline definition**  **(6mmd group)** | **Baseline definition**  **(non-6mmd group)** | **Follow-up period** | **Special considerations**  **for follow-up** |
| --- | --- | --- | --- | --- |
| **Trial 1** | First 6MMD visit within trial period | First clinic visits within trial period | Up to 36 months | 36-month follow-up allowed for 12 or 24-month retention analysis |
| **Trial 2** |  |  | Up to 36 months |  |
| **Trial 3** |  |  | 24 months | 24-month follow-up allowed for 12 retention analysis only based on the administrative censoring |
| **Trial 4** |  |  | 24 months |  |
| Note: The unit of measure is trial-client. Baseline was defined as the first 6MMD visit for individuals in the 6MMD group and the first clinic visit for individuals in the non-6MMD group within each trial period. Follow-up periods vary by trial based on administrative censoring, allowing up to 36 months of follow-up for Trials 1–2 for 24 months retention assessment and 24-month follow-up for Trial 1-4 for 12-month retention analysis. | | | | |

**S2 Table. Characteristics of** **trial-clients eligible for 6MMD in the study in Malawi**

| **Variable** | **Never on 6MMD** | **Ever on 6MMD** | **Total** |
| --- | --- | --- | --- |
| **6MMD status** | (N = 187659) | (N = 76507) | (N = 264166) |
| **Median (IQR) Age in years at first eligible trial** | 35.0 (28.0, 42.0) | 39.0 (32.0, 46.0) | 36.0 (29.0, 43.0) |
| **Age group** |  |  |  |
| 18-34 | 89451 (77.6%) | 25817 (22.4%) | 115268 (100.0%) |
| 35-44 | 63095 (68.8%) | 28647 (31.2%) | 91742 (100.0%) |
| 45-54 | 23657 (61.7%) | 14667 (38.3%) | 38324 (100.0%) |
| 55+ | 11456 (60.8%) | 7376 (39.2%) | 18832 (100.0%) |
| **Gender** |  |  |  |
| Male | 55494 (64.6%) | 30348 (35.4%) | 85842 (100.0%) |
| Female | 132165 (74.1%) | 46159 (25.9%) | 178324 (100.0%) |
| **Duration on ART at first eligible trial** |  |  |  |
| <=12 months | 37231 (71.0%) | 15182 (29.0%) | 52413 (100.0%) |
| 13-24 months | 33939 (72.2%) | 13068 (27.8%) | 47007 (100.0%) |
| 25-36 months | 26263 (70.8%) | 10818 (29.2%) | 37081 (100.0%) |
| 37-48 months | 22794 (70.9%) | 9374 (29.1%) | 32168 (100.0%) |
| >48 months | 67432 (70.6%) | 28065 (29.4%) | 95497 (100.0%) |
| **Facility type** |  |  |  |
| PHC | 91486 (67.6%) | 43769 (32.4%) | 135255 (100.0%) |
| Hospital | 96173 (74.6%) | 32738 (25.4%) | 128911 (100.0%) |
| **Region** |  |  |  |
| Central | 59195 (73.0%) | 21949 (27.0%) | 81144 (100.0%) |
| North | 21049 (72.8%) | 7868 (27.2%) | 28917 (100.0%) |
| South | 107415 (69.7%) | 46690 (30.3%) | 154105 (100.0%) |
| **WHO clinical staging at ART start** |  |  |  |
| WHO stage I/II | 151374 (71.5%) | 60434 (28.5%) | 211808 (100.0%) |
| WHO Stage III/IV | 32292 (68.9%) | 14582 (31.1%) | 46874 (100.0%) |
| Unkown | 3993 (72.8%) | 1491 (27.2%) | 5484 (100.0%) |

**S3 Table. Crude RD and aRD of 12-month retention in care among trial-clients in trial 1-4**

| **Variable** | **Trial 1 (N = 101,979)** | | | | **Trial 2 (N = 68,730)** | | | | **Trial 3 (N = 51,948)** | | | | **Trial 4 (N = 41,509)** | | | |
| --- | --- | --- | --- | --- | --- | --- | --- | --- | --- | --- | --- | --- | --- | --- | --- | --- |
|  | **(N = 9,041)** | **(N = 92,938)** |  |  | **(N = 7,090)** | **(N = 61,640)** |  |  | **(N = 6,137)** | **(N = 45,811)** |  |  | **(N = 6,988)** | **(N = 34,521)** |  |  |
|  | **Not retained** | **Retained** | **Crude RD** | **aRD** | **Not retained** | **Retained** | **Crude RD** | **aRD** | **Not retained** | **Retained** | **Crude RD** | **aRD** | **Not retained** | **Retained** | **Crude RD** | **aRD** |
| **Treatment dispensing interval** |  |  |  |  |  |  |  |  |  |  |  |  |  |  |  |  |
| Non-6MMD | 7 | 43,413 (89.1%) | Reference | Reference | 4661 (11.7%) | 35156 (88.3%) | Reference | Reference | 4027 (13.2%) | 26472 (86.8%) | Reference | Reference | 4393 (16.8%) | 21825 (83.2%) | Reference | Reference |
| 6MMD | 3,732 (7.0%) | 49,525 (93.0%) | 3.9% (3.5%-4.2%) | 3.6% (3.2%-4.0%) | 2429 (8.4%) | 26484 (91.6%) | 3.3% (2.9%-3.8%) | 2.8% (2.3%-3.3%) | 2110 (9.8%) | 19339 (90.2%) | 3.4% (2.8%-3.9%) | 3.1% (2.5%-3.7%) | 2595 (17.0%) | 12696 (83.0%) | -0.2% (-1.0%-0.5%) | 0.0% (-0.8%-0.8%) |
| **Age in years** |  |  |  |  |  |  |  |  |  |  |  |  |  |  |  |  |
| 18-34 | 4,122 (11.4%) | 31,926 (88.6%) | Reference | Reference | 3725 (12.4%) | 26204 (87.6%) | Reference | Reference | 3288 (13.3%) | 21450 (86.7%) | Reference | Reference | 3739 (17.9%) | 17097 (82.1%) | Reference | Reference |
| 35-44 | 2,997 (7.9%) | 35,085 (92.1%) | 3.6% (3.1%-4.0%) | 3.4% (2.9%-3.8%) | 2205 (9.2%) | 21870 (90.8%) | 3.3% (2.8%-3.8%) | 3.2% (2.7%-3.8%) | 1928 (11.0%) | 15600 (89.0%) | 2.3% (1.7%-2.9%) | 2.5% (1.9%-3.2%) | 2197 (16.0%) | 11499 (84.0%) | 1.9% (1.1%-2.7%) | 3.0% (2.1%-3.8%) |
| 45-54 | 1,167 (6.3%) | 1,7261 (93.7%) | 5.1% (4.6%-5.6%) | 5.1% (4.6%-5.6%) | 766 (7.6%) | 9262 (92.4%) | 4.8% (4.2%-5.5%) | 5.2% (4.5%-5.9%) | 623 (9.3%) | 6090 (90.7%) | 4.0% (3.2%-4.8%) | 4.9% (4.0%-5.8%) | 734 (15.2%) | 4107 (84.8%) | 2.8% (1.7%-3.9%) | 5.3% (4.0%-6.5%) |
| 55+ | 755 (8.0%) | 8,666 (92.0%) | 3.4% (2.8%-4.1%) | 3.4% (2.8%-4.1%) | 394 (8.4%) | 4304 (91.6%) | 4.1% (3.2%-4.9%) | 4.6% (3.6%-5.5%) | 298 (10.0%) | 2671 (90.0%) | 3.3% (2.1%-4.4%) | 4.3% (3.1%-5.5%) | 318 (14.9%) | 1818 (85.1%) | 3.1% (1.5%-4.7%) | 5.4% (3.7%-7.1%) |
| **Sex** |  |  |  |  |  |  |  |  |  |  |  |  |  |  |  |  |
| Male | 3,996 (11.0%) | 32,178 (89.0%) | Reference | Reference | 2670 (12.6%) | 18507 (87.4%) | Reference | Reference | 2322 (14.7%) | 13521 (85.3%) | Reference | Reference | 2588 (20.5%) | 10060 (79.5%) | Reference | Reference |
| Female | 5,045 (7.7%) | 60,760 (92.3%) | 3.4% (3.0%-3.8%) | 4.3% (3.9%-4.7%) | 4420 (9.3%) | 43133 (90.7%) | 3.3% (2.8%-3.8%) | 4.4% (3.8%-4.9%) | 3815 (10.6%) | 32290 (89.4%) | 4.1% (3.5%-4.7%) | 5.1% (4.4%-5.8%) | 4400 (15.2%) | 24461 (84.8%) | 5.2% (4.4%-6.0%) | 6.2% (5.3%-7.1%) |
| **Duration on ART** |  |  |  |  |  |  |  |  |  |  |  |  |  |  |  |  |
| <=12 months | 1,109 (13.9%) | 6,887 (86.1%) | Reference | Reference | 1325 (13.7%) | 8369 (86.3%) | Reference | Reference | 1350 (14.7%) | 7836 (85.3%) | Reference | Reference | 1572 (17.7%) | 7298 (82.3%) | Reference | Reference |
| 13-24 months | 2,062 (11.9%) | 15,314 (88.1%) | 2.0% (1.1%-2.9%) | 1.8% (0.9%-2.7%) | 1553 (13.3%) | 10154 (86.7%) | 0.4% (-0.5%-1.3%) | 0.5% (-0.4%-1.5%) | 1415 (14.1%) | 8601 (85.9%) | 0.6% (-0.4%-1.6%) | 0.6% (-0.4%-1.6%) | 1680 (18.7%) | 7290 (81.3%) | -1.0% (-2.1%-0.1%) | -1.2% (-2.4%-0.0%) |
| 25-36 months | 1,749 (10.3%) | 15,245 (89.7%) | 3.6% (2.7%-4.5%) | 3.1% (2.3%-4.0%) | 1173 (11.1%) | 9361 (88.9%) | 2.5% (1.6%-3.4%) | 2.4% (1.4%-3.3%) | 925 (12.7%) | 6373 (87.3%) | 2.0% (1.0%-3.1%) | 1.9% (0.8%-2.9%) | 977 (17.9%) | 4466 (82.1%) | -0.2% (-1.5%-1.1%) | -0.7% (-2.1%-0.6%) |
| 37-48 months | 1,279 (8.6%) | 13,616 (91.4%) | 5.3% (4.4%-6.2%) | 4.6% (3.7%-5.5%) | 905 (10.3%) | 7876 (89.7%) | 3.4% (2.4%-4.3%) | 3.1% (2.2%-4.1%) | 713 (12.2%) | 5142 (87.8%) | 2.5% (1.4%-3.6%) | 2.5% (1.3%-3.6%) | 681 (16.2%) | 3510 (83.8%) | 1.5% (0.1%-2.8%) | 1.0% (-0.4%-2.4%) |
| >48 months | 2,842 (6.4%) | 41,876 (93.6%) | 7.5% (6.7%-8.3%) | 6.4% (5.6%-7.2%) | 2134 (7.6%) | 25880 (92.4%) | 6.1% (5.3%-6.8%) | 5.2% (4.4%-6.0%) | 1734 (8.9%) | 17859 (91.1%) | 5.9% (5.0%-6.7%) | 5.6% (4.7%-6.5%) | 2078 (14.8%) | 11957 (85.2%) | 2.9% (1.9%-3.9%) | 3.2% (2.1%-4.3%) |
| **Facility type** |  |  |  |  |  |  |  |  |  |  |  |  |  |  |  |  |
| PHC | 4,557 (8.4%) | 49,560 (91.6%) | 0.9% (0.6%-1.3%) | 0.9% (0.5%-1.2%) | 3445 (10.1%) | 30632 (89.9%) | 0.4% (0.0%-0.9%) | 0.7% (0.2%-1.2%) | 2939 (11.7%) | 22266 (88.3%) | 0.3% (-0.3%-0.9%) | 1.5% (0.9%-2.1%) | 3573 (16.3%) | 18283 (83.7%) | 1.0% (0.3%-1.8%) | 2.7% (1.9%-3.5%) |
| Hospital | 4,484 (9.4%) | 43,378 (90.6%) | Reference | Reference | 3645 (10.5%) | 31008 (89.5%) | Reference | Reference | 3198 (12.0%) | 23545 (88.0%) | Reference | Reference | 3415 (17.4%) | 16238 (82.6%) | Reference | Reference |
| **Region** |  |  |  |  |  |  |  |  |  |  |  |  |  |  |  |  |
| Central | 2,564 (8.3%) | 28,409 (91.7%) | Reference | Reference | 2221 (10.4%) | 19223 (89.6%) | Reference | Reference | 2231 (13.7%) | 14006 (86.3%) | Reference | Reference | 2844 (22.8%) | 9646 (77.2%) | Reference | Reference |
| North | 1,322 (11.8%) | 9,867 (88.2%) | -3.5% (-4.2%-2.9%) | -3.4% (-4.1%-2.6%) | 1000 (13.5%) | 6427 (86.5%) | -3.1% (-4.0%-2.2%) | -2.8% (-3.7%-1.8%) | 770 (13.4%) | 4974 (86.6%) | 0.3% (-0.7%-1.4%) | 1.1% (0.0%-2.2%) | 718 (15.8%) | 3839 (84.2%) | 7.0% (5.7%-8.3%) | 8.2% (6.9%-9.6%) |
| South | 5,155 (8.6%) | 54,662 (91.4%) | -0.3% (-0.7%-0.0%) | 0.6% (0.2%-1.0%) | 3869 (9.7%) | 35990 (90.3%) | 0.7% (0.2%-1.2%) | 1.6% (1.1%-2.1%) | 3136 (10.5%) | 26831 (89.5%) | 3.3% (2.6%-3.9%) | 4.5% (3.8%-5.1%) | 3426 (14.0%) | 21036 (86.0%) | 8.8% (7.9%-9.6%) | 9.8% (8.9%-10.6%) |
| **WHO clinical staging at ART start** |  |  |  |  |  |  |  |  |  |  |  |  |  |  |  |  |
| WHO stage I/II | 7,256 (9.3%) | 71,038 (90.7%) | Reference | Reference | 5922 (10.7%) | 49350 (89.3%) | Reference | Reference | 5204 (12.1%) | 37666 (87.9%) | Reference | Reference | 5976 (16.9%) | 29396 (83.1%) | Reference | Reference |
| WHO Stage III/IV | 1,722 (7.9%) | 20,170 (92.1%) | 1.4% (1.0%-1.8%) | -0.3% (-0.8%-0.1%) | 1096 (9.1%) | 10917 (90.9%) | 1.6% (1.0%-2.2%) | -0.2% (-0.8%-0.4%) | 843 (10.9%) | 6884 (89.1%) | 1.2% (0.5%-2.0%) | -0.3% (-1.2%-0.5%) | 897 (17.1%) | 4345 (82.9%) | -0.2% (-1.3%-0.9%) | -0.5% (-1.6%-0.7%) |
| Unknown | 63 (3.5%) | 1,730 (96.5%) |  |  | 72 (5.0%) | 1373 (95.0%) |  |  | 90 (6.7%) | 1261 (93.3%) | 0 | 0 | 115 (12.8%) | 780 (87.2%) |  |  |
| Notes: Values are presented as n/N (% retained) for each trial (Trial 1-4). Risk differences (RD) show percentage point differences in 12-month retention between individuals on 6MMD versus never on 6MMD, with 95% confidence intervals (CIs). Adjusted RDs are adjusted forage at trial entry, sex, ART duration at trial entry, facility type, WHO stage, and region. Abbreviations: ART: antiretroviral therapy; CI: confidence interval; 6MMD: six-month dispensing; RD: risk difference; WHO: World Health Organization | | | | | | | | | | | | | | | | |

**S4 Table. Crude and aRD of 24-month retention in care among trial-clients in trial 1-2**

|  | **Trial 1 (N = 101979)** | | | | **Trial 2 (N = 68730)** | | | |
| --- | --- | --- | --- | --- | --- | --- | --- | --- |
|  | **(N = 9041)** | **(N = 92938)** |  |  |  | **(N = 7090)** | **(N = 61640)** |  |
| **Variable** | **Not retained** | **Retained** | **Crude RD** | **aRD** | **Not retained** | **Retained** | **Crude RD** | **aRD** |
| Non-6MMD | 8,251 (16.9%) | 40,471 (83.1%) | Reference | Reference | 87,54 (22.0%) | 31,063 (78.0%) | Reference | Reference |
| 6MMD | 6,819 (12.8%) | 46,438 (87.2%) | 4.13% (3.00%-5.26%) | 3.42% (2.96%-3.88%) | 6,099 (21.1%) | 22,814 (78.9%) | 0.89% (-0.45%-2.23%) | -0.12% (-0.77%-0.53%) |
| **Age in years** |  |  |  |  |  |  |  |  |
| 18-34 | 6585 (18.3%) | 29,463 (81.7%) | Reference | Reference | 7,431 (24.8%) | 22,498 (75.2%) | Reference | Reference |
| 35-44 | 5179 (13.6%) | 32,903 (86.4%) | 4.67% (3.35%-5.99%) | 4.53% (3.98%-5.08%) | 4,788 (19.9%) | 19287 (80.1%) | 4.94% (3.44%-6.44%) | 5.24% (4.50%-5.98%) |
| 45-54 | 2067 (11.2%) | 16,361 (88.8%) | 7.05% (5.40%-8.70%) | 7.15% (6.50%-7.80%) | 1,782 (17.8%) | 8,246 (82.2%) | 7.06% (5.03%-9.09%) | 8.22% (7.25%-9.19%) |
| 55+ | 1239 (13.2%) | 8,182 (86.8%) | 5.12% (3.02%-7.22%) | 5.21% (4.37%-6.04%) | 852 (18.1%) | 3,846 (81.9%) | 6.69% (3.93%-9.46%) | 7.65% (6.36%-8.94%) |
| **Gender** |  |  |  |  |  |  |  |  |
| Male | 6,266 (17.3%) | 29,908 (82.7%) | Reference | Reference | 5,175 (24.4%) | 16,002 (75.6%) | Reference | Reference |
| Female | 8,804 (13.4%) | 57,001 (86.6%) | 3.94% (2.77%-5.12%) | 5.07% (4.57%-5.57%) | 9,678 (20.4%) | 37,875 (79.6%) | 4.08% (2.67%-5.50%) | 5.40% (4.66%-6.14%) |
| **Duration on ART** |  |  |  |  |  |  |  |  |
| <=12 months | 1,654 (20.7%) | 6,342 (79.3%) | Reference | Reference | 2,500 (25.8%) | 7,194 (74.2%) | Reference | Reference |
| 13-24 months | 3,266 (18.8%) | 14,110 (81.2%) | 1.89% (-0.48%-4.26%) | 1.66% (0.59%-2.73%) | 2,957 (25.3%) | 8,750 (74.7%) | 0.53% (-1.79%-2.85%) | 0.51% (-0.68%-1.70%) |
| 25-36 months | 2,855 (16.8%) | 14,139 (83.2%) | 3.89% (1.50%-6.27%) | 3.58% (2.52%-4.64%) | 2,448 (23.2%) | 8,086 (76.8%) | 2.55% (0.15%-4.95%) | 2.30% (1.10%-3.51%) |
| 37-48 months | 2,141 (14.4%) | 12,754 (85.6%) | 6.31% (3.86%-8.76%) | 5.78% (4.71%-6.84%) | 1,913 (21.8%) | 6,868 (78.2%) | 4.00% (1.48%-6.53%) | 3.84% (2.59%-5.09%) |
| >48 months | 5,154 (11.5%) | 39,564 (88.5%) | 9.16% (7.02%-11.30%) | 8.34% (7.37%-9.30%) | 5,035 (18.0%) | 22,979 (82.0%) | 7.82% (5.80%-9.83%) | 7.70% (6.66%-8.74%) |
| **Facility type** |  |  |  |  |  |  |  |  |
| PHC | 7,413 (13.7%) | 46,704 (86.3%) | 2.30% (1.17%-3.43%) | 2.97% (2.49%-3.45%) | 7,387 (21.7%) | 26,690 (78.3%) | -0.13% (-1.46%-1.19%) | 2.21% (1.54%-2.88%) |
| Hospital | 7,657 (16.0%) | 40,205 (84.0%) | Reference | Reference | 7,466 (21.5%) | 27,187 (78.5%) | Reference | Reference |
| **Region** |  |  |  |  |  |  |  |  |
| Central | 4,880 (15.8%) | 26,093 (84.2%) | Reference | Reference | 5,877 (27.4%) | 15,567 (72.6%) | Reference | Reference |
| North | 1,904 (17.0%) | 9285 (83.0%) | -1.26% (-3.23%-0.71%) | 0.01% (-0.85%-0.88%) | 1,657 (22.3%) | 5,770 (77.7%) | 5.10% (2.79%-7.40%) | 6.08% (4.89%-7.26%) |
| South | 8,286 (13.9%) | 51,531 (86.1%) | 1.90% (0.64%-3.17%) | 3.48% (2.97%-4.00%) | 7,319 (18.4%) | 32,540 (81.6%) | 9.04% (7.60%-10.50%) | 10.90% (10.20%-11.70%) |
| **WHO clinical staging at ART start** |  |  |  |  |  |  |  |  |
| WHO stage I/II | 11,996 (15.3%) | 66,298 (84.7%) | Reference | Reference | 12,222 (22.1%) | 43,050 (77.9%) | Reference | Reference |
| WHO Stage III/IV | 2,958 (13.5%) | 18,934 (86.5%) | 1.81% (0.42%-3.20%) | -0.00467 | 2,413 (20.1%) | 96,00 (79.9%) | 2.03% (0.27%-3.79%) | -0.45% (-1.31%-0.41%) |
| Unknown | 116 (6.5%) | 1,677 (93.5%) |  |  | 218 (15.1%) | 1,227 (84.9%) |  |  |
| **Notes:** Values are presented as n/N (% retained) for each trial (Trial 1-2). Risk differences (RD) show percentage point differences in 24-month retention between individuals on 6MMD versus never on 6MMD, with 95% confidence intervals (CIs). Adjusted RDs are adjusted forage at trial entry, sex, ART duration at trial entry, facility type, WHO stage, and region. ART: antiretroviral therapy; CI: confidence interval; 6MMD: six-month dispensing; RD: risk difference; WHO: World Health Organization | | | | | | | | |
